# Psychometric properties of the Alimetry Gut-Brain Wellbeing Survey-II: A revised, unidirectional mental health scale for patients with chronic gastroduodenal symptoms

**DOI:** 10.64898/2026.09.30.26364444

**Authors:** Mikaela Law, Isabella Pickering, Gabriel Schamberg, Charlotte Daker, Armen Gharibans, Greg O’Grady, Christopher N. Andrews, Stefan Calder

## Abstract

**Objective:** The Alimetry Gut-Brain Wellbeing (AGBW) Survey is a validated 10-item scale developed to assess mental health in patients with chronic gastroduodenal symptoms. While the original scale demonstrated strong psychometric properties overall, as part of an ongoing commitment to continuous instrument optimization, this study evaluated a revised, unidirectional version of the scale (AGBW-II) to enhance measurement precision.

**Methods:** The psychometric properties of the revised AGBW-II were evaluated via an anonymous online questionnaire in a sample of 161 patients with chronic gastroduodenal symptoms. Score distributions, factor analysis, reliability, and validity were analyzed and compared with data from the validation study of the original scale (n= 311).

**Results:** The AGBW-II total and subscale scores demonstrated robust reliability and validity, outperforming the original AGBW Survey. Notable improvements in the reliability and validity of the stress subscale were observed following item rewording. Confirmatory factor analysis verified the three-factor model separating the depression, stress, and anxiety subscales, with the AGBW-II showing improved model fit indices.

**Conclusions:** The revised, unidirectional AGBW-II demonstrated improved reliability and validity, while maintaining the underlying psychometric properties of the original scale. Consequently, the AGBW-II provides a reliable, valid, and brief mental health assessment for patients with chronic gastroduodenal symptoms.

## Introduction

Chronic gastroduodenal symptoms constitute a major socioeconomic and clinical issue, affecting upwards of 10% of the population globally, with both their prevalence and associated healthcare costs continuing to climb [1–4]. Psychological comorbidities, such as depression, anxiety, and stress, are highly prevalent in these patients through a bidirectional relationship with the gut-brain axis, and play a critical role in modulating symptom severity, clinical compliance, and overall quality of life [5–8]. Consequently, psychological assessments are commonly regarded as a critical part of the standard care for these patients [9,10].

The Alimetry Gut-Brain Wellbeing (AGBW) Survey was developed as a brief screening tool to assess mental health in patients with chronic gastroduodenal symptoms [11]. In alignment with best practices in patient-reported outcome development, feedback from real-world use highlighted an opportunity to further refine the scale. The original 10-item scale included two reverse-worded items (items 5 and 7). While mixed-direction item-phrasing is traditionally used to reduce response bias, standardizing all items into a unidirectional format reduces cognitive processing load and improves measurement clarity.

To improve clinical utility, data quality, and item comprehension, the scale was modified into a unidirectional format, phrasing all items in the negative direction (AGBW-II). The primary objective of this study was to examine the psychometric properties of the AGBW-II and validate it against the original version to investigate how this iterative optimization could enhance reliability, without compromising the scale’s measurement capabilities.

## Methods

### Scale Modification

The original AGBW Survey [11] consists of a patient preface, 10 closed-ended questions asking patients to rate how often they have felt or behaved in a certain way over the last two weeks on a scale of (none of the time) to 4 (all the time), and an optional open-ended item. A total score and three distinct subscales can be calculated by totalling items 1-4 (depression subscale), items 5-7 (stress subscale), and items 8-10 (anxiety subscale). Higher scores indicate worse mental health. Items 5 and 7 (both in the stress subscale) were written in a positive frame and therefore need to be reverse-coded before score calculations.

For the AGBW-II, items 5 and 7 were modified slightly to a unidirectional format, in line with the rest of the scale items, by adding the word “not” to both items (see Table 1). Face validity of the newly worded items was assessed with a group of clinical gastroenterologists and specialized researchers in the area of psychogastroenterology. The full AGBW-II is found in the Supplementary Material.

**Table 1.** Modifications made to the AGBW-II wording, with added words highlighted in bold.

| Item Number | Original Wording | New Wording |
| --- | --- | --- |
| 5 | Over the last 2 weeks, how often have you felt like you could cope with the challenges in your life? | 1. Over the last 2 weeks, how often have you felt like you could **not** cope with the challenges in your life? |
| 7 | Over the last 2 weeks, how often have you felt like things were going well for you? | 1. Over the last 2 weeks, how often have you felt like things were **not** going well for you? |

### Procedure

To allow for direct comparisons between the old and new scales, the procedures, measures, eligibility criteria, and data analysis for this validation study were identical to those of the original scale validation, which involved a sample of 311 patients [11]. Psychometric validation of the scale was conducted using an anonymous, cross-sectional survey of patients with chronic gastroduodenal symptoms. All patients provided informed consent. The anonymous survey was completed online via Qualtrics (Qualtrics, Provo, UT) and involved demographic questions and a battery of psychological questionnaires, including the AGBW-II, presented in a randomized order. Responses were collected between June and July 2026. Ethical approval was granted by the Auckland Health Research Ethics Committee Application (AH25798), and the trial was pre-registered at ANZCTR.org.au (ACTRN12623000385640).

### Sample

Patients were recruited globally via convenience sampling through social media and were included if they were over 18 years old and able to speak, read, and write fluently in English. Patients also had to meet the Rome IV symptom criteria [12] and/or have a self-reported clinical diagnosis for at least one of the following conditions: gastroparesis, functional dyspepsia, chronic nausea and vomiting syndrome, cyclic vomiting syndrome, rumination syndrome, cannabinoid hyperemesis syndrome, or a belching disorder. Patients with self-induced vomiting or an eating disorder were excluded.

### Measures

#### Demographics

Participants provided basic demographics and self-reported whether they had ever been diagnosed with a mental health issue.

#### Psychometrics

The following psychometrics were measured to assess convergent validity: the Patient Health Questionnaire 9 (PHQ-9) [13], the Generalized Anxiety Disorder 7 (GAD-7) [14], the Perceived Stress Scale 4 (PSS-4) [15], the Depression Anxiety and Stress Scale 21 (DASS-21) [16], and the Kessler Psychological Distress Scale (K-10) [17]. The Big Five Inventory (BFI) extraversion subscale [18] and the Emotion Regulation Questionnaire (ERQ) [19] were measured to assess divergent validity. Lastly, concurrent validity was assessed using the Patient Assessment of Upper Gastrointestinal Disorders-Quality of Life (PAGI-QOL) [20].

### Statistical Analysis

Data were analyzed using IBM SPSS Statistics v29. A *p*-value of .05 was considered statistically significant. To assess score-level equivalence, descriptive comparisons were made for the subscale and total score means and distributions between the AGBW-II and the original version of the scale, as detailed in Law et al. [11]. Additionally, independent-samples t-tests comparing the subscale and total scores of the old and new versions of the scale were conducted; a lack of statistical significance indicated score-level equivalence.

Reliability, validity, and confirmatory factor analysis of the AGBW-II subscales and total score were assessed using the same predetermined acceptability criteria used in the validation study for the AGBW Survey and compared to the original study results (see Law et al. [11] for the full statistical analysis plan and original results).

## Results

### Sample Characteristics

A total of 161 participants completed the online survey (mean age= 37.03 years, *SD*= 14.14, range= 18-76 years). Demographics are provided in Table 2.

**Table 2.** Demographic characteristics of the survey respondents.

|  | *n* (%) |
| --- | --- |
| Gender |  |
| Female | 122 (76%) |
| Male | 31 (19%) |
| Gender diverse | 8 (5%) |
| Region |  |
| North America | 84 (52%) |
| Europe | 41 (25%) |
| Oceania | 33 (21%) |
| Asia | 2 (1%) |
| Central America | 1 (1%) |
| Rome IV Criteria<sup>a</sup> |  |
| Functional dyspepsia | 138 (86%) |
| Chronic nausea and vomiting syndrome | 125 (78%) |
| Cyclic vomiting syndrome | 28 (17%) |
| Cannabinoid hyperemesis syndrome | 4 (1%) |
| Rumination syndrome | 57 (35%) |
| Belching disorder | 66 (41%) |
| Diagnosis of gastroparesis | 63 (39%) |
| Previous mental health diagnoses |  |
| Depression | 86 (53%) |
| Anxiety | 105 (65%) |
| None | 42 (26%) |

*Notes:* *n* = number of respondents in that category; % = percentage of respondents in that category; <sup>a</sup> Rome IV criteria diagnosis is based on meeting the symptom criteria only; respondents can meet more than one Rome IV diagnosis.

### Descriptive Statistics

Similar to the original scale, all three subscales and the total score were normally distributed (see Table 3). There were slight decreases in the mean scores for the stress subscale and total score compared to the original scale. However, these deviations were only small, with independent samples t-tests between the old and new scores showing non-significance (total score: *mean difference=* -0.50, *p*= .561; depression subscale score: *mean difference=* -0.13, *p*= .718; stress subscale score: *mean difference=* -0.45, *p*= .108; anxiety subscale score: *mean difference=* 0.08, *p*= .799).

**Table 3.** Descriptive statistics of the AGBW-II subscales and total scores.

| Scale | *M* | *SD* | Range | Skewness | Kurtosis | $\alpha$ |
| --- | --- | --- | --- | --- | --- | --- |
| Depression subscale | 7.98 | 3.79 | 0-16 | 0.04 | -0.68 | .80 |
| Stress subscale | 5.64 | 3.33 | 0-12 | 0.10 | -0.79 | .90 |
| Anxiety subscale | 5.29 | 3.43 | 0-12 | 0.32 | -0.93 | .87 |
| Total score | 18.90 | 9.46 | 1-40 | 0.23 | -0.77 | .92 |

Notes: *M*= mean; *SD*= standard deviation, $\alpha$ = Cronbach's alpha coefficient.

### Confirmatory Factor Analysis

Confirmatory factor analysis revealed that the three-factor model, splitting the depression, stress, and anxiety subscales, had a good model of fit, with superior model fit indices compared to the original version of the scale (*χ*^*2*^_(32)_= 53.45, *p=* .010, *χ*^*2*^*/df*= 1.67, NFI= .95, CFI= .98, TLI= .97, SRMR= .03, RMSEA= .07 [.03-.09]).

### Reliability

The Cronbach’s *α* coefficients demonstrated good to excellent internal consistency reliability for the AGBW-II total score and subscale scores (Table 3). In particular, the stress subscale score (containing the revised items) increased from acceptable (*α*= 0.72) to excellent (*α*= 0.90), while the other scores maintained their high reliability. Inter-item correlations (*r*= .33-.76) and corrected item-total correlations (*r*= .55-.79) also improved relative to the original scale, primarily owing to greater reliability of the revised items 5 and 7 in the stress scale.

### Validity

The unidirectional version of the scale successfully maintained acceptable convergent, divergent, and concurrent validity, satisfying all predetermined acceptability criteria (Table 4). Additionally, the convergent and concurrent validity of the stress subscale score was superior to that of the original scale.

**Table 4.**
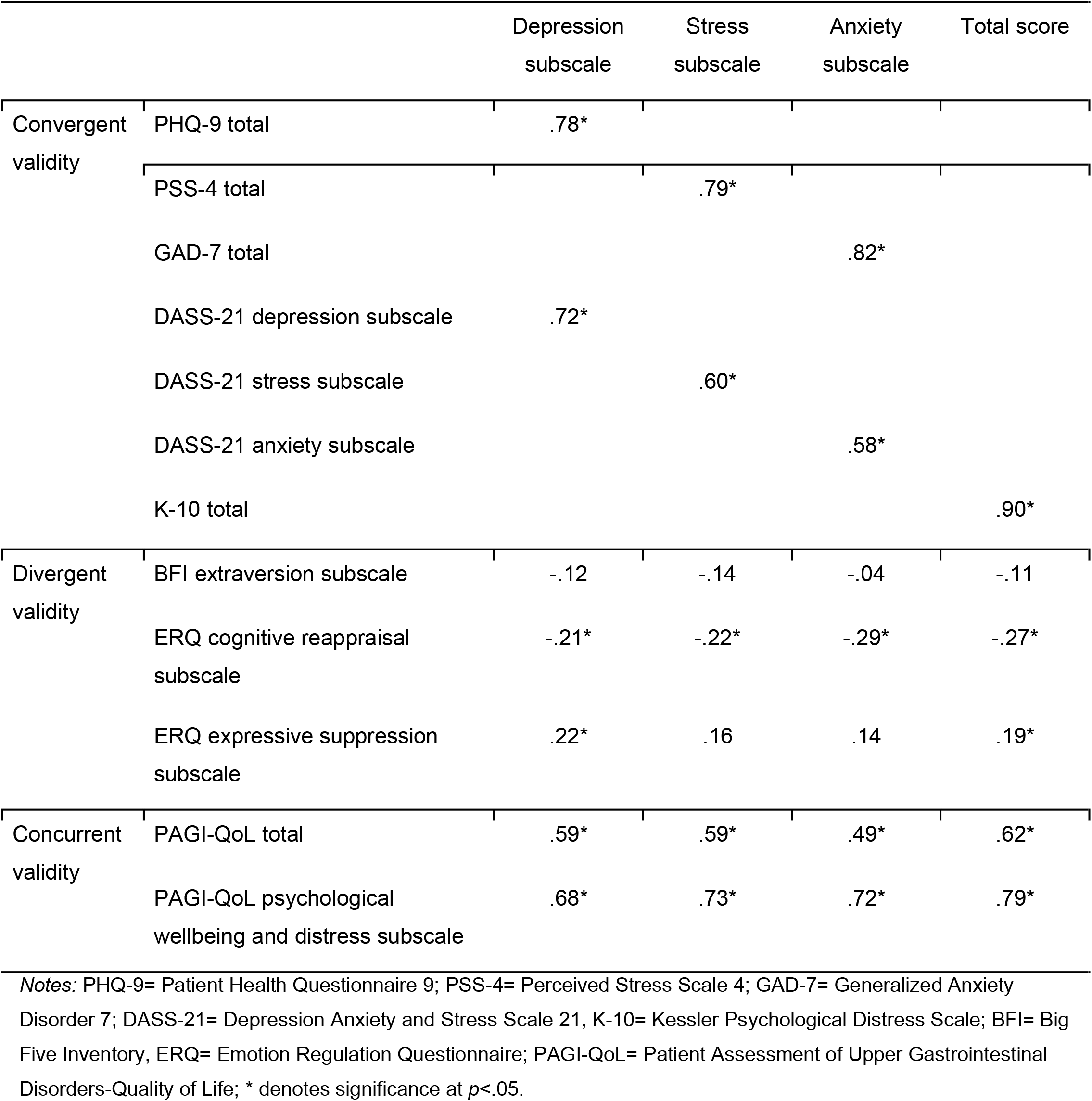
Pearson correlation coefficients between the AGBW-II scores and the comparative questionnaires used for validity testing.

|  |  | Depression subscale | Stress subscale | Anxiety subscale | Total score |
| --- | --- | --- | --- | --- | --- |
| Convergent validity | PHQ-9 total | .78* |  |  |  |
|  | PSS-4 total |  | .79* |  |  |
|  | GAD-7 total |  |  | .82* |  |
|  | DASS-21 depression subscale | .72* |  |  |  |
|  | DASS-21 stress subscale |  | .60* |  |  |
|  | DASS-21 anxiety subscale |  |  | .58* |  |
|  | K-10 total |  |  |  | .90* |
| Divergent validity | BFI extraversion subscale | -.12 | -.14 | -.04 | -.11 |
|  | ERQ cognitive reappraisal subscale | -.21* | -.22* | -.29* | -.27* |
|  | ERQ expressive suppression subscale | .22* | .16 | .14 | .19* |
| Concurrent validity | PAGI-QoL total | .59* | .59* | .49* | .62* |
|  | PAGI-QoL psychological wellbeing and distress subscale | .68* | .73* | .72* | .79* |

*Notes:* PHQ-9= Patient Health Questionnaire 9; PSS-4= Perceived Stress Scale 4; GAD-7= Generalized Anxiety Disorder 7; DASS-21= Depression Anxiety and Stress Scale 21, K-10= Kessler Psychological Distress Scale; BFI= Big Five Inventory, ERQ= Emotion Regulation Questionnaire; PAGI-QoL= Patient Assessment of Upper Gastrointestinal Disorders-Quality of Life; \* denotes significance at $p < .05$ .

Additionally, known groups validity was also confirmed; those who self-reported a previous mental health diagnosis scored, on average, significantly higher on all three subscales and the total score than those who did not have a previous mental health diagnosis, with large effect sizes (all *p*< .001, *d*= .70-.81).

## Discussion

This research demonstrated that modifying the AGBW into a unidirectional format (AGBW-II) improved scale comprehension, reliability, and validity, without compromising its underlying psychometric properties. Rephrasing the two reverse-coded items in the stress scale appeared to reduce cognitive processing load and input errors, supporting improved scale comprehension.

As expected, the most substantial psychometric gains were observed within the stress subscale, where the item rewording occurred. Specifically, the internal consistency of the stress subscale improved from acceptable to excellent reliability, accompanied by increases in convergent and concurrent validity. Furthermore, confirmatory factor analysis demonstrated that the foundational three-factor structure separating the depression, stress, and anxiety subscales was superior, demonstrating strong model fit across the revised scale items. Additionally, the total and subscale scores remained normally distributed across the sample, with only small, non-significant shifts in the mean scores for the stress subscale and total score observed. Rather than indicating scale drift, this minor shift likely reflects enhanced clarity and a reduction in artificial measurement noise previously associated with mixed-direction phrasing.

Overall, these findings confirm that the AGBW-II is highly reliable and valid, providing confidence in data collection consistency. Consequently, the AGBW-II provides direct practical benefits for both clinical and research settings. Ambiguous or confusing item wording can artificially skew a patient’s score, risking the over- or under-reporting of their mental health [21,22]. By removing this source of error, the AGBW-II ensures clinicians and researchers receive an accurate assessment of a patient’s true mental health. Furthermore, standardizing the instrument into a unidirectional layout reduces the cognitive burden required for completion. This supports ease of data collection, especially in busy clinic settings, and accommodates patients experiencing severe symptomology, which may heighten cognitive strain. Ultimately, the AGBW-II supports the collection of high-quality, reliable psychometric data that can be combined with objective physiological assessments, such as body surface gastric mapping [23]. This integrated approach helps guide comprehensive, multidisciplinary care, including psychological referrals and targeted behavioral interventions.

Despite the demonstrated psychometric improvements, the risks of unidirectional scale design must be acknowledged. Transitioning to a unidirectional design inherently increases susceptibility to other response biases, such as straight-lining or social desirability bias, which, in clinical practice, could result in false negatives and the under-reporting of mental health concerns [22]. While such bias was not evident in our anonymous research sample, it remains a risk in real-world clinical settings. This is especially a concern among patients worried that acknowledging mental health concerns could lead clinicians to dismiss their gastrointestinal symptoms as purely psychological [24]. Additionally, this study evaluated cross-cohort differences between two separate patient samples, rather than utilizing a repeated-measures design, which may have limited the conclusions.

In summary, revising the AGBW to the unidirectional AGBW-II optimized reliability and validity, while maintaining its psychometric properties. The AGBW-II provides a brief, valid, and highly reliable mental health assessment tailored specifically for patients with chronic gastroduodenal symptoms. It provides gastroenterologists and psychologists with actionable psychometric data to complement objective physiological testing and inform multidisciplinary care in routine clinical practice.

## Data Availability

Deidentified data will be made available upon reasonable request to the corresponding author.

## Notes

**Conflicts of interest:** GOG and AG are Directors of the University of Auckland spin-out company Alimetry Ltd. ML, GS, CD, CNA, and SC are members of Alimetry Ltd. GOG, AG, GS, and SC hold intellectual property and grants in gastric electrophysiology. The remaining authors have no relevant conflicts to declare.

### Competing Interest Statement

GOG and AG are Directors of the University of Auckland spin-out company Alimetry Ltd. ML, GS, CD, CNA, and SC are members of Alimetry Ltd. GOG, AG, GS, and SC hold intellectual property and grants in gastric electrophysiology. The remaining authors have no relevant conflicts to declare.

### Clinical Trial

ACTRN12623000385640

### Author Declarations

All patients provided informed consent. Ethical approval was granted by the Auckland Health Research Ethics Committee Application (AH25798), and the trial was pre-registered at ANZCTR.org.au (ACTRN12623000385640).

