## Supplementary Material for "Psychometric properties of the Alimetry Gut-Brain Wellbeing Survey-II: A revised, unidirectional mental health scale for patients with chronic gastroduodenal symptoms"

The following 10 questions will ask you about your mental wellbeing over the past 2 weeks.

Research has shown that stomach symptoms and mental wellbeing can affect each other through the gut-brain axis, a physical connection between the stomach and brain.

These questions will help your clinician better understand your mental wellbeing in connection with your symptoms and stomach's activity. This allows for a more holistic understanding of your condition, enabling a more personalised management plan.

Please note these questions cannot be used to diagnose you with a mental health condition and are not intended to attribute your symptoms to your mental health. Your answers are confidential and will be seen by your referring clinician in your test report.

- ☐ Decline wellbeing survey

  Feel free to comment on why you have chosen not to answer these questions (optional)

  ______________________________

- ☐ Continue

**1. Over the last 2 weeks, how often have you felt a reduced interest in things that usually bring you enjoyment?**

| None of the time | A little of the time | Some of the time | Most of the time | All of the time |
|---|---|---|---|---|
| ☐ | ☐ | ☐ | ☐ | ☐ |

**2. Over the last 2 weeks, how often have you felt sad, depressed, or unhappy?**

| None of the time | A little of the time | Some of the time | Most of the time | All of the time |
|---|---|---|---|---|
| ☐ | ☐ | ☐ | ☐ | ☐ |

**3. Over the last 2 weeks, how often have you felt tired, fatigued, or lacking in energy, for no good reason?**

| None of the time | A little of the time | Some of the time | Most of the time | All of the time |
|---|---|---|---|---|
| ☐ | ☐ | ☐ | ☐ | ☐ |

**4. Over the last 2 weeks, how often have you found thinking, staying focused, or making decisions difficult?**

| None of the time | A little of the time | Some of the time | Most of the time | All of the time |
|---|---|---|---|---|
| ☐ | ☐ | ☐ | ☐ | ☐ |

© Alimetry Ltd., 2026. Please contact for any licensing requests

5. **Over the last 2 weeks, how often have you felt like you could not cope with the challenges in your life?**

| None of the time | A little of the time | Some of the time | Most of the time | All of the time |
|---|---|---|---|---|
| ☐ | ☐ | ☐ | ☐ | ☐ |

6. **Over the last 2 weeks, how often have you felt like the important things in your life were out of your control?**

| None of the time | A little of the time | Some of the time | Most of the time | All of the time |
|---|---|---|---|---|
| ☐ | ☐ | ☐ | ☐ | ☐ |

7. **Over the last 2 weeks, how often have you felt like things were not going well for you?**

| None of the time | A little of the time | Some of the time | Most of the time | All of the time |
|---|---|---|---|---|
| ☐ | ☐ | ☐ | ☐ | ☐ |

8. **Over the last 2 weeks, how often have you felt anxious, nervous, or unable to relax?**

| None of the time | A little of the time | Some of the time | Most of the time | All of the time |
|---|---|---|---|---|
| ☐ | ☐ | ☐ | ☐ | ☐ |

9. **Over the last 2 weeks, how often have you found it hard to stop worrying about things?**

| None of the time | A little of the time | Some of the time | Most of the time | All of the time |
|---|---|---|---|---|
| ☐ | ☐ | ☐ | ☐ | ☐ |

10. **Over the last 2 weeks, how often have you felt scared or afraid as if something bad might happen, for no good reason?**

| None of the time | A little of the time | Some of the time | Most of the time | All of the time |
|---|---|---|---|---|
| ☐ | ☐ | ☐ | ☐ | ☐ |

**Please provide any further comments about your answers in the box below that you would like your clinician to be aware of (optional)**

© Alimetry Ltd., 2026. Please contact for any licensing requests
